# Assessing the acceptability and feasibility of proactive community case management: A qualitative study in Chadiza District, Zambia

**DOI:** 10.1101/2025.07.08.25331096

**Authors:** Bupe M. Kabamba, Chabu C. Kangale, Melody N. Simataa, Marie-Reine I. Rutagwera, Travis Porter, John M. Miller, Ellen Leah Ferriss, Caroline Phiri-Chibawe, Maximillian Musunse, Patrick Nyendwa, Viennah Kapenda, Paul Psychas, Julie R. Gutman, Adam Bennett, Ignatius Banda, Sampa Chitambala-Otiono, Busiku Hamainza, Julie I. Thwing

## Abstract

Zambia has implemented passive malaria community case management (CCM), during which symptomatic people seek care at a community health worker’s (CHW) abode, since 2011. In 2021, the National Malaria Elimination Program implemented a two-arm cluster-randomized controlled trial to measure the impact of proactive CCM (ProCCM) compared to routine passive CCM on malaria incidence and prevalence in Chadiza District, Eastern Province. ProCCM is a strategy of regular visits by CHWs to all households in a community to identify people with malaria symptoms, provide rapid diagnostic testing, and treat those with malaria with artemisinin combination therapy. We conducted a qualitative midline assessment of this trial to describe the feasibility and acceptability of ProCCM. Key informant interviews (KIIs) were conducted with CHWs provincial and district staff, health facility staff, and community leaders, while focus group discussions were conducted with community members from six ProCCM and six passive CCM clusters. All respondent groups preferred ProCCM to passive CCM, reporting that it reduces distance travelled to seek care and provides care to people with challenges accessing it. Most CHWs and health facility staff perceived the ProCCM arm to have less malaria compared to the passive arm. Six provincial, district, and health facility staff and five CHWs felt that ProCCM was onerous for CHWs, necessitating compensation. Four health facility staff reported supervising ProCCM CHWs more frequently than before the trial, which two saw as an additional, albeit manageable, time commitment. District and health facility staff noted an increase in commodities consumption due to ProCCM. Other challenges included long distances covered by CHWs during visits and the impact of the farming cycle on both households’ and CHWs’ availability for visits. The findings suggest that ProCCM is both feasible to implement and acceptable to communities but requires additional time and travel for CHWs.

## Introduction

Malaria is among the leading causes of death globally, with sub-Saharan Africa bearing most of the morbidity and mortality burden. Globally, between 2022 and 2023, malaria cases increased by 11 million from 252 million to 263 million and deaths decreased slightly from 600,000 to 597,000 [1]. Most of these cases and deaths occurred in sub-Saharan African countries such as Zambia. Zambia accounts for 1.4 % of the global malaria case and death burden [1]. Improving access to and use of health services for prompt and effective case management of malaria has been a persistent challenge. In 2011, the National Malaria Elimination Program (NMEP) introduced integrated community case management (iCCM), in which community health workers (CHWs) are trained to offer a menu of basic services focused on maternal and child health. By 2021, about 40% of confirmed malaria cases were detected by CHWs nationally [2]. Despite this, some malaria cases remain undiagnosed and untreated, contributing to ongoing transmission and increased morbidity. For example, nationwide, less than half of febrile children under 5 years of age were promptly taken to a trained medical provider in 2021 [3].

Proactive community case management (ProCCM) of malaria has been used to fill in the gaps left by iCCM. In iCCM, CHWs provide passive malaria community case management (mCCM), in which community members with malaria symptoms may seek care at the CHW’s home. In ProCCM, CHWs visit households within their catchment area every one to two weeks to detect people with malaria symptoms, offer diagnosis by RDT, and treat those with positive results with an ACT, in addition to being available between household visits to provide malaria diagnosis and treatment services as in routine passive mCCM. This serves to increase the proportion of the community who receive timely diagnosis and treatment [4] and strengthens ties between CHW and community members.

A non-randomized cluster-controlled trial of ProCCM vs. routine mCCM conducted in Senegal found a 16-fold reduction of confirmed symptomatic malaria infection in the ProCCM arm at endline [5]. Senegal has since scaled ProCCM to over 2000 communities in higher transmission districts [6]. In addition, ProCCM has recently been studied in various settings in Madagascar [7], Mali [8], and Uganda [9]. However, none of these trials were conducted in a setting with the combination of a pre-existing, highly functional mCCM program in addition to health facility case management, indoor residual spraying (IRS), and high coverage of insecticide treated nets (ITNs). While Senegal has published a description of the scale-up, others have not published data related to acceptance and feasibility.

We conducted a trial of ProCCM in Chadiza district, Zambia, in a setting with a strong mCCM program and high IRS and ITN coverage, to assess the impact on incidence and prevalence of malaria. As part of this trial, we conducted a qualitative study to assess the acceptance and feasibility of ProCCM among Ministry of Health (MoH) staff, CHWs, community leaders, and community members. ProCCM was both feasible to implement and acceptable to communities but required additional resources, including time commitment from CHWs and remuneration for their work.

## Materials and methods

### Study design

This was a cross-sectional qualitative assessment conducted as a midline survey of a two-arm cluster randomized controlled trial measuring the impact of ProCCM compared to routine passive CCM on malaria incidence and prevalence in Chadiza District, Eastern Province, Zambia. (Details of the RCT are presented elsewhere [10].)

### Study setting

Chadiza, a predominantly agrarian district located in the Eastern province of Zambia, is a seasonal and moderate transmission setting with a malaria incidence of 200 to 499 cases per 1000 persons per year and parasite prevalence of 10% to 20%. Scale up of malaria community case management began in 2019; by 2021, Chadiza had 161 CHWs with a CHW to population ratio of 1:500 based on the national malaria program guidelines. Community utilization of the CHWs is high, with 60% to 70% of all malaria cases reported by the district diagnosed and treated by CHWs at community level [2]. The district also received high coverage of vector control interventions (IRS and ITN). In 2021, more than 90% and 30% of the population were protected by IRS and ITN, respectively [11].

### Study context

We conducted a cluster randomized controlled trial of ProCCM vs. routine passive mCCM from 2021 to 2023, beginning with a baseline household survey in April-May 2021 and ending with an endline household survey in April-May 2023 (Fig 1). After the baseline survey, CHWs conducted a household registration exercise during which household members’ names, age, and sex were recorded in the CommCare application [12]. Thereafter, 66 CHW catchment areas (clusters) were randomized 1:1 into the intervention and control arms, with 33 clusters per arm (Fig 2), ensuring that the 2 arms were balanced with respect to prevalence. Both intervention and control arm CHWs were selected from among the 161 iCCM trained CHWs providing routine passive mCCM. Each CHW is attached to a health facility. As such, CHWs may be assigned to undertake other health care services in the facility.

**Fig 1.**
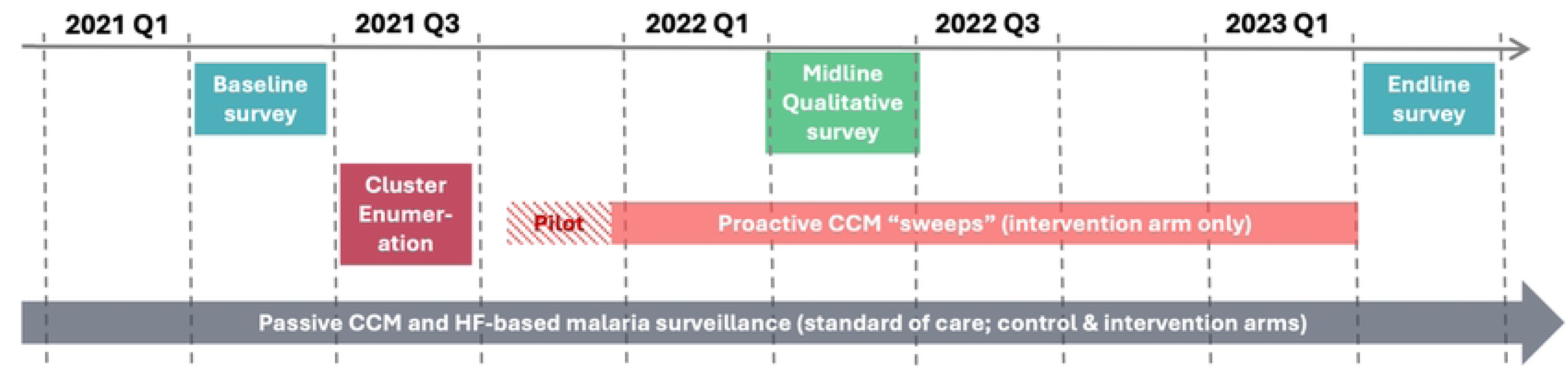
ProCCM study timeline.

**Fig 2.**
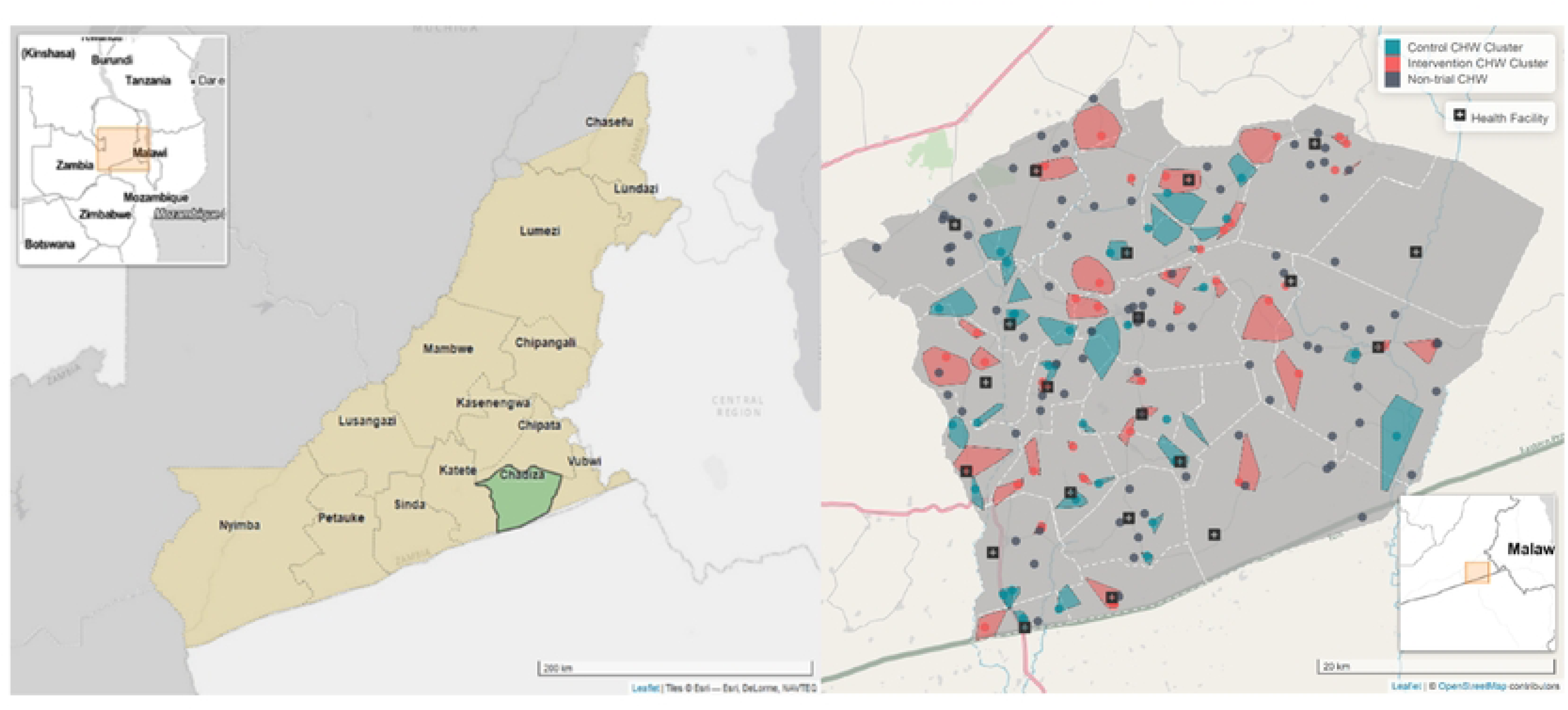
**Study site showing CHW catchment areas, or clusters, receiving routine CCM plus proCCM (the intervention) in red and routine CCM only in blue.**

In both arms, the target population was all household members. Control arm clusters implemented routine passive mCCM while intervention arm clusters implemented ProCCM (which includes access to routine passive mCCM). The ProCCM strategy used weekly visits by CHWs to all households in their assigned catchment area to identify people with malaria symptoms, offer diagnostic testing, and treat those with positive tests. Implementation of ProCCM began in December 2021 and ended in May 2023 (at the time of the endline survey).

During each ProCCM visit, CHWs used the household listing in the Commcare app to record whether anyone in the household had malaria symptoms and, if anyone was found with symptoms, to select that household member from the pre-populated list and record responses to questions regarding symptoms, the RDT result, whether ACT treatment was given, and whether a referral was made to a health facility. The app used these data to generate quantitative data reports that included the number of people during each weekly visit (1) with symptoms of malaria, (2) tested, (3) who tested positive for malaria, (4) referred to a health facility, as well as the number of visits that each CHW conducted to each household and the approximate time they spent at each household. Throughout the study, CHWs in both arms received support through regular meetings with the research and DHO team ensuring availability of RDTs and ACTs, supervision, and provision of enablers such as personal protective equipment and bicycles.

Prior to initiation of the intervention, communities in both arms were sensitized on the study and the difference between the two approaches. Periodic and responsive community stakeholder engagement was implemented throughout the study period. The qualitative study was conducted in July in both intervention and control arm clusters.

### Data collection

Key informant interviews (KIIs) and focus groups discussions (FGDs) with participants recruited from July through August, 2022, were conducted by MoH staff from other districts within Eastern Province who were trained in qualitative data collection methods. The assessment was conducted in 12 of the 66 study clusters: six in the intervention arm and six in the control arm. The clusters were selected using criteria that included selecting one cluster far from a health facility, one cluster close to a health facility, two clusters with a wide geographic dispersion of houses, and two clusters with low dispersion of houses. Key informant interviews (KIIs) were conducted with 12 CHWs and 33 community leaders while 26 focus group discussions (FGDs) were conducted with community members. Each FGD included 6-10 participants. Other key informants were 6 provincial and district staff who manage the health system at sub-national level and 9 health facility (HF) staff who supervise CHWs. KIIs were conducted in the preferred language of the respondent, either English or Chichewa, while all FGDs were conducted in Chichewa (Table 1).

**Table 1.**
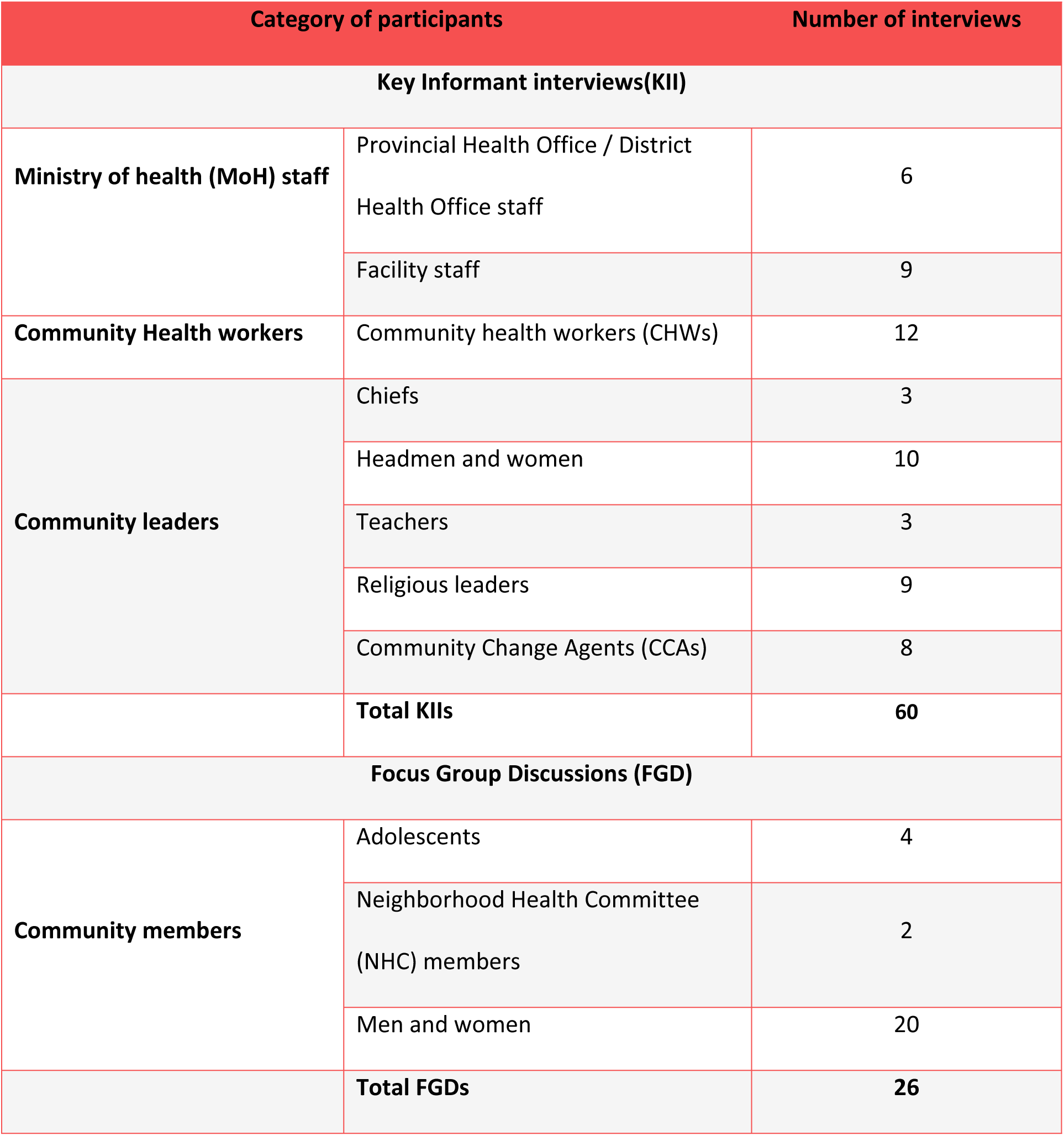
Study participant categories.

| Category of participants |  | Number of interviews |
| --- | --- | --- |
| **Key Informant interviews(KII)** |  |  |
| **Ministry of health (MoH) staff** | Provincial Health Office / District Health Office staff | 6 |
|  | Facility staff | 9 |
| **Community Health workers** | Community health workers (CHWs) | 12 |
| **Community leaders** | Chiefs | 3 |
|  | Headmen and women | 10 |
|  | Teachers | 3 |
|  | Religious leaders | 9 |
|  | Community Change Agents (CCAs) | 8 |
|  | **Total KIIs** | **60** |
| **Focus Group Discussions (FGD)** |  |  |
| **Community members** | Adolescents | 4 |
|  | Neighborhood Health Committee (NHC) members | 2 |
|  | Men and women | 20 |
|  | **Total FGDs** | **26** |

All FGDs and KIIs were done using a semi-structured interview guide adapted for each participant category. The guides were structured enough to guide the interview and flexible to allow for probing when needed.

Feasibility was defined as the plausibility and practicability of CHWs conducting household visits once every week year-round. It was assessed among CHWs in the intervention arm; in addition, health facility staff were asked whether they were able to provide supervision to the CHWs in the study and if frequency of supervision changed. To assess feasibility, the study examined three domains: a) Labor demands: whether CHWs visited all households in their cluster once every week (assessed using Commcare data), still had time to attend to their day-to-day economic and social activities, and their perceptions of conducting ProCCM; b) Availability of malaria commodities: whether CHWs always had RDTs and ACTs and c) CHW Supervision: Perceived change in frequency of supervision compared to before the study and whether it was manageable.

Acceptance was measured by first assessing the CommCare app data for CHW visit completion rate. A completed visit was defined as a household visit where the CHW was able to find out if there was anyone with malaria symptoms, irrespective of whether anyone in the household reported malaria symptoms. Additionally, acceptance was assessed from the FGDs and KIIs by coding positive views about the acceptability of ProCCM, including expressions of satisfaction, intent to continue the use of ProCCM, perceived appropriateness, and perceived positive effects on stakeholders [13]. Responses were categorized into three themes: a) whether community members accepted CHWs visiting their homes every week; b) community members’ perceptions of CHWs household visits, and c) community members preferred frequency of household visits.

### Data analysis

A code book was deductively developed based on the objectives of the study from a priori codes from the interview guides. For completeness, additional codes were generated inductively and discussed by the coding team when further information was found in the transcripts. The team developed a code book using a thematic approach and coded all the transcripts using QDA miner version 2.0.9 software [14]. Coding segments were used to identify quotes, participant categories, and disaggregation between the two study arms.

### Ethical considerations

The study was reviewed and approved by Zambian ERES Converge Research Ethics Committee (Ref no: 2020-Jan-003), the National Health Research Authority of Zambia, PATH’s Research Ethics Committee (PATH REC), the US Centers for Disease Control and Prevention IRB (7278), and Western Copernicus Group (WCG) IRB (IRB Protocol #20210568) to ensure adherence to research ethical standards. Written informed consent was obtained from all participants aged 18 years and older, written assent was obtained from all adolescent respondents aged 15-17 years. The purpose of the study, the time that the interview would take, benefits, risks, and confidentiality were explained to all the participants.

Confidentiality was emphasized and maintained throughout the interview. During FGDs participants were not referred to by their names, instead, each participant was assigned a number, and they were called to express themselves by that number. The interviewees were informed that the interviews would be recorded and that the conversations would not be disclosed to anyone. Recorded interviews were stored in a secure cloud-based content management system called Box file [15]. Once they were transcribed, the files were locked with a password on the Box file.

## Results

### Feasibility

#### Did community health workers visit all households once every week?

Between December 2021, when the household visits began, and July 2022, the time of qualitative data collection (an eight-month period), CHWs made 129,806 of 156,264 (83.1%) expected visits, and . conducted an average of 26 visits per household over 32 weeks. Figure 3 is an example of CHWs’ weekly visits tracked through CommCare. The total proportion of expected visits conducted varied weekly among CHWs, though three-fourths of CHWs visited at least 75% of eligible households in the average week. The average number of households in intervention clusters was 153 with a range of 61 to 309.

**Fig 3.**
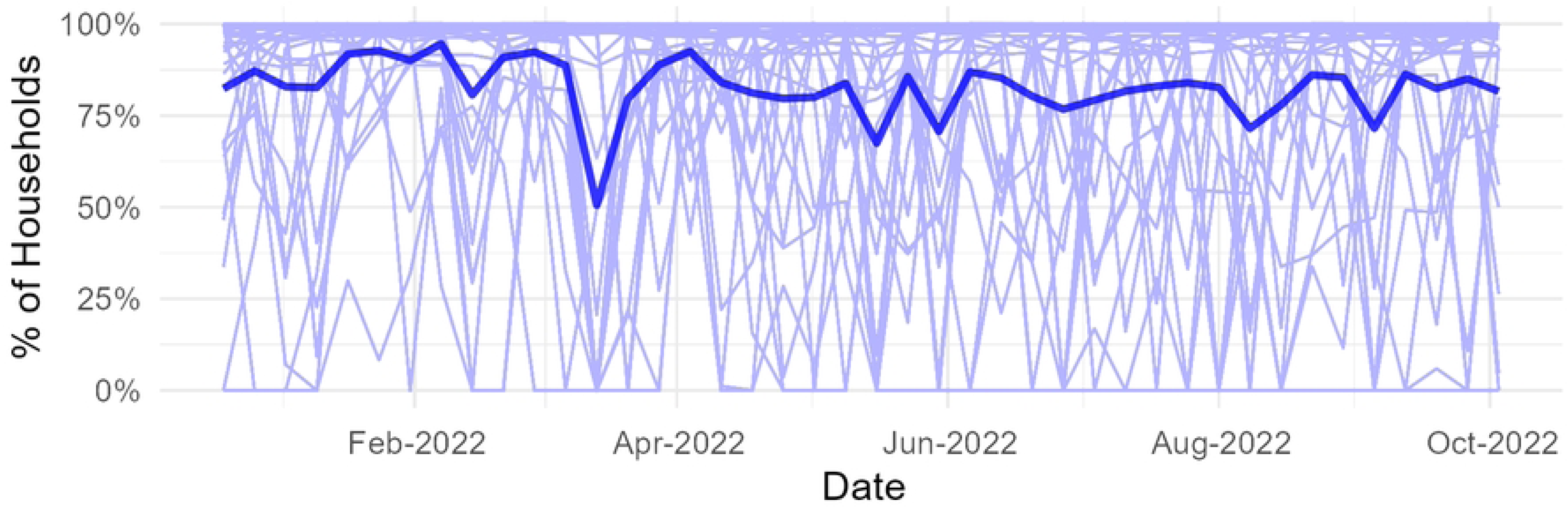
Weekly percentage of eligible household visits conducted by CHWs. Each faint blue line represents an individual CHWs’ weekly attempted visits to households and shows that the concentration of the visit is close to 100%, while the thick blue line is for one CHW showing how their weekly visits fluctuated over time.

Therefore, on average CHWs were expected to make 153 weekly visits. Every proactive (intervention) CHW visited at least 49 households in the average week; three-fourths of CHWs visited at least 87 households in the average week. CHWs spent an average of three days each week (range 2-5 days) to reach all households in the cluster.

CHWs’ opinions of the practicability of conducting weekly visits were mixed. Half of them found doing visits easy while the other half expressed various concerns. Those that found ProCCM feasible attributed it to having a clear weekly plan that enabled them to apportion time between ProCCM and their personal business and to their commitment to helping the community overcome malaria.

> *“I have developed a timetable which I follow. I have separated time for ProCCM activities and for personal work. I usually do my personal work in the morning and ProCCM activities in the afternoon when I am assured to find people in their households. I spend four days. I start on Monday and finish on Thursday. Within this period, I manage to finish all the households in my cluster. I rest over the weekend and begin on Monday again.” KII, Intervention arm CHW*

The other half expressed concerns about ProCCM. They found the exercise an inconvenience and the workload challenging because the study competed against their personal social and family activities.

> *“…before ProCCM it was good because I was just waiting for people at home but now whether you like it or not every week you have to leave home to test people in their households. So, it’s a different way that has reduced the programs I do at home. In the past I had a lot of time to do housework but now I have less time to do housework because most of the time is spent doing ProCCM work” KII, Intervention arm CHW*

Ministry of Health (facility and DHO staff) also questioned the feasibility of weekly visits by CHWs. Many felt that the workload was onerous for CHWs.

> *“…this program [ProCCM] needs someone who is committed to work because it is so involving…most of them complain [that] they don’t have time for other activities they just focus on the program [ProCCM] and there is need to be incentivizing [them].” KII, Facility staff*

Similarly, most community members and leaders shared this view. They perceived ProCCM as full-time work warranting compensation given the time CHWs invested each week visiting households.

> *“A CHW to leaving his personal work and go door to door is difficult for him to take care of his family. This can only work if the government, donors or community allocated something for him as a salary. So that he is just doing that work without attending to his personal duties.” FGD, Men control arm*

> *“We know that these people [CHWs] may need an allowance because they are moving a very long distance. Sometimes they are covering between 5 −10km.” KII, Traditional leader*

Distance covered between houses by CHWs was an important factor in the manageability of conducting ProCCM. Dispersion of households varied among clusters with the most dispersed cluster having a centroid distance of 1.4kms and the least having 0.3 kms. As such, the need for transport to conduct household visits was almost universally mentioned among the intervention CHWs--even among the CHWs who found ProCCM manageable.

> *“ Working in the ProCCM is harder as it requires following people in their households, and this requires reliable transport.” KII, Intervention arm CHW*

> *“There is need to compensate the CHWs who are doing ProCCM especially [with] the means of transport like bicycles as they sacrifice their time and effort serving the community.” KII, Intervention arm CHW*

#### Availability of malaria commodities: Did CHWs always have RDTs and ACTs?

CHWs and MoH staff were asked if they ever experienced commodity stockouts during the ProCCM trial. Most reported not experiencing any stockouts, stating that malaria commodities (RDTs and ACTs) were always available.

> *“We never stocked out in all the commodities for malaria – antimalarial, RDTs – at least we have had all those commodities throughout since the study started.” KII, Health facility staff*

In cases where health facilities reported an anticipated stock out, the district pharmacist worked with health facilities to redistribute commodities from those overstocked to those at risk of running out and later balanced the required number of commodities in all facilities to ensure sustained availability.

> *“We have had some facilities that would [be] stocked out maybe in about 2 to 3 days. They communicate and then you find that there is another facility that still has stock maybe for a month or so, so [we] would get some stock from that facility and provide it to the other facility as we wait for the central level to provide us with our commodities. So, yes through that we’ve maintained stock in our facilities” DHO Staff*

Generally, CHWs in both arms were adequately stocked throughout the trial period (Fig 4). By the end of the month, on average, CHWs had a minimum of 25 RDTs as balance on hand. This stock was adequate to start the new month even if they were not replenished by the facility at the beginning of the new month, a strategy deliberately agreed upon by the study team, provincial and district MoH staff.

**Fig 4.**
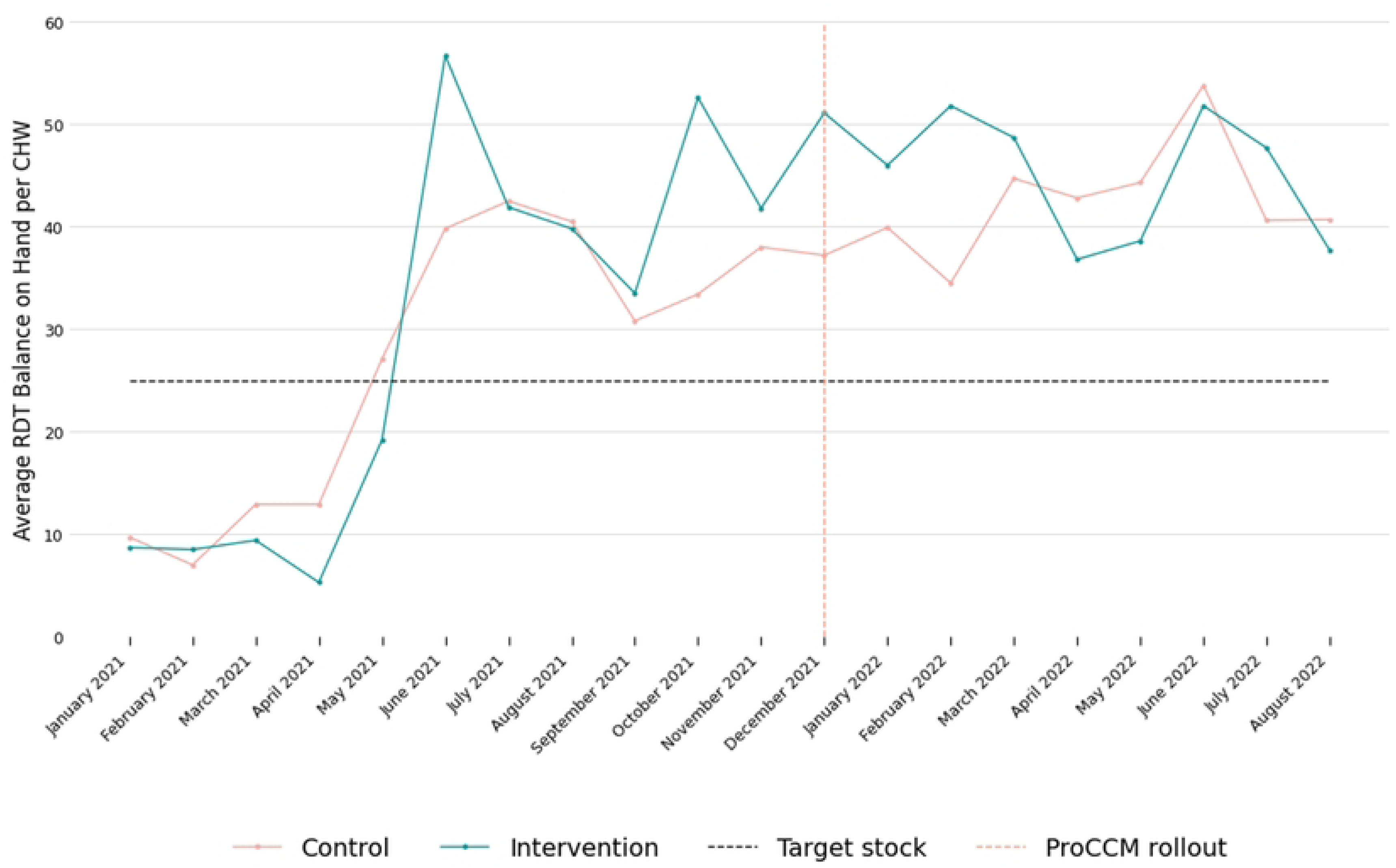
**Average monthly RDT balance on hand per CHW.**

Intervention arm CHWs were asked whether they perceived an increase in the consumption of malaria commodities during the trial, many of whom thought there was not.

> *“There isn’t a change, because at times when you’re working in ProCCM, you might go out as I have mentioned but not find anyone sick. You might find that you will not use RDTs the whole week because you won’t find anyone with malaria signs, so you won’t test anyone. So, materials that we used in the past and now, there’s little change.” KII, Intervention arm CHW*

The few intervention-arm CHWs who perceived an increase in consumption of commodities since the beginning of the trial expressed having to request commodities more frequently than before. None of the CHWs expressed a reduction in supply of malaria commodities.

> *“…nowadays we run out [of commodities] fast compared to the time before the ProACT started. This is because of household visits that I do once per week.” – KII, Intervention arm CHW*

The Ministry of Health staff, however, thought that ProCCM had increased consumption of RDTs and ACTs.

> *“We are seeing now almost triple the consumption rate following the start of the ProACT study.” DHO staff*

Using testing rate and positivity rate as a proxy measure for consumption, quantitative data showed that consumption of RDTs in the intervention arm was higher than control (Fig 5).

**Fig 5.**
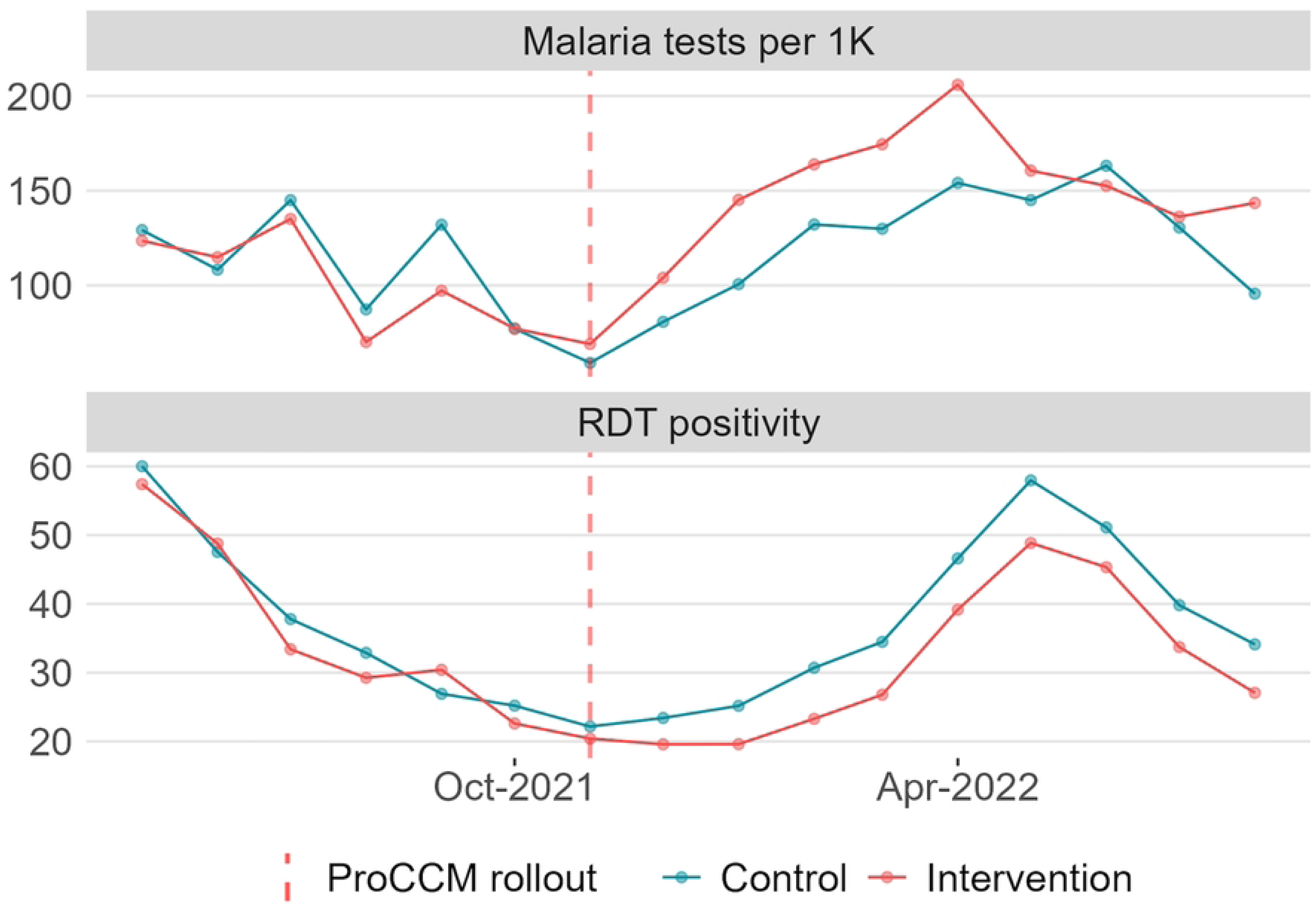
**RDT per 1000 population done**.

#### CHW Supervision: Did it change and was it manageable?

The NMEP recommends that to sustain quality of community case management, CHWs should be supervised by health facility staff trained as CHW supervisors. During supervision visits, using the NMEP’s standard supervision checklist, supervisors check whether CHWs have adequate malaria commodities and other supplies, and whether CHWs are abiding by prescribed community case management standards. In KIIs, CHWs and supervisors (health facility staff) were asked whether CHWs were supervised and whether ProCCM increased the number of contacts between supervisors and CHWs to understand the impact ProCCM would have on supervisors’ time. CHWs in both arms reported being supervised by health facility staff during the study, with half of intervention arm CHWs reporting an increase in frequency of supervision.

> *“Before ProCCM started we were rarely visited compared to now.” CHW intervention arm*

Similarly, most health facility staff supervising CHWs in the intervention arm reported increasing the frequency of their supervision. This increased their workload but was mostly perceived to be manageable.

> *“[Supervision was] maybe quarterly or so. Now the frequency of visiting them has also improved.” KII, Health facility staff*

> *“It [ProCCM supervision] affects my workload but not very much because that is part of my work, working in the field with CHWs. However, due to lack of human resources in our health facilities, less attention has been given to other activities because most of the time I’m busy with ProCCM activities.” – KII, Health facility staff*

### Acceptance

#### Did community members accept CHWs visiting their homes?

Weekly visits by CHWs to households entailed synchronizing the timing of a CHW visit to the availability of household members. Members of the household needed to be present at the time of the visit for the visit to be successful and for the community members to benefit from the service. CHWs’ household visits were classified as expected visits, attempted visits, and completed visits. Expected visits was the total number of visits that a CHWs should have conducted dependent on the number of households in their cluster, an attempted visit was when a CHW visited the household whether or not they found anyone at the household to respond to the visit, while a completed visit was one in which a CHW visited the household and found people who were able to report if there was anyone with malaria symptoms, regardless of whether anyone actually reported malaria symptoms. From December 2021 to July 2022, at the time of the qualitative assessment, CHWs made a combined 129,806 attempted visits to households out of which 122,157 (94.1%) were completed. CHWs were able to find someone at home to respond to the questionnaire for nearly all (94.1%) households during their weekly visits (Fig 6).

**Fig 6.**
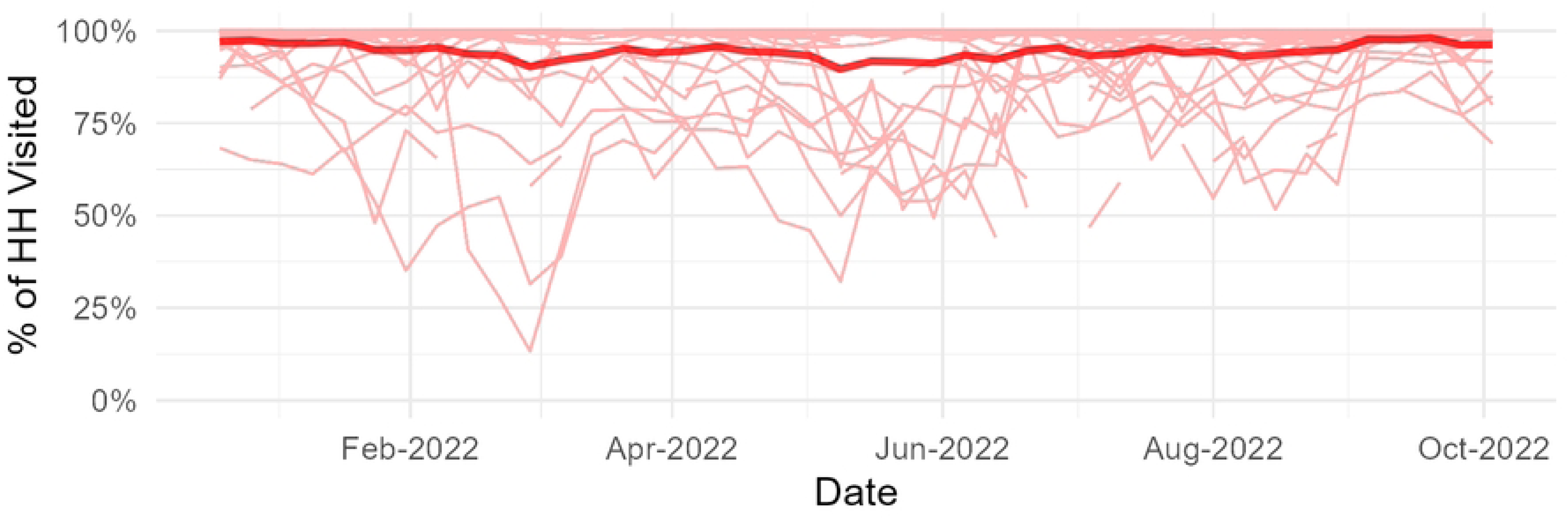
Percentage of attempted visits completed overall and by cluster. The light red lines show the percentage of attempted visits that were completed for individual CHWs while the solid deep red line shows the completion rate by one CHW, both showing a high completion rate close to 100%.

CHWs found a total of 9,763 people with malaria symptoms, out of which they tested 9,745 (99.8%). Of those tested, 2,692 tested positive, representing a positivity rate of 27.6%. The high proportion oof completed visits and tests done during these visits indicate very high acceptance of ProCCM by community members because community members not only allowed CHWs to visit households, but they also agreed to be tested for malaria when they were symptomatic.

##### Community members’ perceptions of CHWs household visits

In FGDs and KIIs, most respondents viewed ProCCM positively, while a few expressed some concerns. Those with positive views noted ProCCM benefitted community members by providing malaria services within their homes.

> *“This idea of visiting door to door is good because there are some of us who are lazy to go to the clinic or to the community health worker. When [the CHW] visits he finds me and tests me he will have helped me even if I felt lazy to go to the hospital in the first place.” FGD, Women, Intervention arm*

For MoH personnel, ProCCM fulfilled the ministry’s goal of bringing health care as close to people’s homes as possible.

> *“What makes it appealing is when community health workers follow people at home and diagnose malaria, then treat [people] in their homes without them going to facilities. It cuts on the distances that one would cover making it easy to reach people.” KII, District Health Office Staff*

> *“That’s what the Ministry of Health wants to do, we want to provide services as close to the families as possible so that they don’t have to move distances.” KII, Provincial Health Office Staff*

##### Community members preferred frequency of household visits

Many respondents preferred at least one visit every week. Others perceived such frequent visits as being taxing on CHWs therefore once a month or less would be more appropriate.

> *“Community health workers should be visiting households at least twice or three times per week” KII, community change agent (CCA), intervention arm.*

> *“I think once a week is okay, because he is also under pressure at his home with people who visit him at his house, and he also visits others. So, I think once weekly is okay because he visits us every week.” FGD, Women, Intervention arm*

> *“The weekly one should be maintained…so that if found sick the treatment is given. Once a week is fine because these people have a lot of other things to do.” FGD, Adolescent female, Control arm*

## Discussion

The study sought to assess whether ProCCM in addition to routine passive CCM is a feasible and acceptable approach in Chadiza, Zambia. Findings suggest that ProCCM is feasible, as CHWs were able to visit most households at the scheduled weekly frequency. Over half of CHWs felt that the ProCCM household visits were quite manageable, often with the help of the scheduling tool introduced early in the implementation period to assist CHWs with planning their time. In contrast to the ProCCM implementation in Senegal, in which CHWs were able to designate one day per week to conduct their visits to every household [4], communities in Zambia were more dispersed, with a higher population.

The alternate strategy of allocating periods of several days appears to have been effective for some CHWs in Zambia.

However, ProCCM visits were perceived as onerous by many informants and took CHWs away from socio-economic activities. This suggests that scaling up ProCCM in Zambia would need a remuneration framework. During the trial (shortly after this assessment), the need to compensate intervention-arm CHWs for ProCCM visits became apparent to the study team, based on the feedback from MoH and CHWs in study review meetings. In response, the study team instituted performance-based remuneration of $22 per month to intervention CHWs reaching at least 75% of all households in their cluster. In addition, CHWs in both arms were provided with enablers such as bicycles, boots, raincoats, phones and phone credit to transmit data. The WHO recommends that remuneration for CHWs be commensurate with demands, complexity, number of hours, training, and roles that they are assigned [16]. Both ProCCM programs that have been scaled or studied on a large scale (Senegal and Mali) have found that it was important to compensate CHWs for their time spent on case management activities. In Senegal [6], CHWs often serve as distributors in campaigns (season al malaria chemoprevention, ITN distribution, deworming and vitamin A supplementation, etc.) and are compensated for that work. The one day of household visits weekly is considered equivalent to a day spent as a campaign distributor for purposes of compensation [5]. In Mali, CHWs receive a monthly salary for providing ProCCM services [8].

Regular and systematic supervision has been shown to improve CHW performance [17]. Although the study did not seek to improve supervision of CHWs by facility staff, it appears to have done so. Both CHWs and facility staff reported increased frequency of supervision. The use of a digital tool may have increased opportunities for supervision. CHW activity submitted to the Commcare server was monitored weekly by the study team. This allowed for a quick way for health facility staff to check on, diagnose, and troubleshoot issues and offer the necessary support to CHWs with performance or reporting issues, such as over or underreporting or repeated visits to particular households. In Mali, the digital tool was used not only to offer high quality supervision, but to help CHWs plan their household visits [18].

Health facility staff are often in charge of supervising CHWs, requiring them to travel distances to communities that may not be accessible by road and taking them away from responsibilities at the health facility. In the routine passive CCM model, supervisors do not often have the opportunity to observe CHW-patient consultations. In the ProCCM model, they can schedule supervision during household visits to maximize opportunities to observe the CHW offering diagnosis and treatment [5]. Supervisors may also play a role in replenishing the CHWs’ stocks of RDTs and ACTs.

This qualitative assessment found that ProCCM was acceptable to community members. This was evident not only in the proportion of visits responded to by households (94.1%) but also in the proportion of people that accepted to be tested when found symptomatic during these visits (99.8%). Synchronizing time of visit by the CHW and availability of household members was crucial. Identifying appropriate times to visit and the CHWs being picked from and by the community they serve was found to be a significant factor in acceptance of door-to-door malaria elimination interventions in Tanzania [19]. Acceptance of ProCCM was largely driven by reduced distance to care, and provision of care to people who otherwise would have not sought care. Door-to-door testing for HIV was acceptable in Malawi for the same reasons [20]. The ministry of health saw ProCCM as an approach that can help achieve its mission of providing healthcare services as close to the family as possible [21].

### Limitations

This study had four main limitations, the first that it was done six months into the study period of 18 months, in July, which is around the end of peak malaria season and harvest season, so it may not account for changes in acceptance and feasibility vis a vis changes in malaria and farming seasons.

Secondly, the interviews were conducted by MOH personnel, which may have potentially introduced a degree of desirability bias in the respondents. Thirdly, while the study included diverse respondents from both arms, the analysis did not compare results between the arms, sex of respondents, or before and after implementing the performance-based remuneration. Lastly, feasibility did not include a cost analysis. A cost-effectiveness analysis could provide valuable insights into feasibility.

## Conclusions

These results suggest that ProCCM is both feasible to implement and acceptable (and indeed desirable) to communities. Over a seven-month period, CHWs conducted >75% of the targeted number of visits to households and provided services to community members in all the households in each cluster. However, several elements are required for the sustained implementation of ProCCM, most criticially, compensation for CHWs that conduct regular household visits. On average, CHWs dedicated half a week to implementing ProCCM, which took time away from their social and economic life. Given the high opportunity cost, it is important to compensate them for their time. Transportation was identified as critical for ProCCM feasibility in this rural context in which communities are widely dispersed. Visiting every household, especially in rural areas, involves covering considerable distances. It is therefore important that CHWs are provided with a bicycle or other form of transportation to successfully carry out ProCCM. Continuous availability of malaria commodities was the third element identified as critical to feasibility, as lack of RDTs and ACTs would negate the purpose of conducting visits and damage both CHW and community confidence in the efficacy of the program, potentially resulting in lower CHW utilization and delayed diagnoses. Chadiza district community members accepted ProCCM as an additional tool in malaria case management at the community level. They perceived ProCCM as bringing health benefits to the community in rural areas where distance to care can be a challenge. Bringing services to the doorstep increased the opportunity to diagnose and treat malaria as early as possible, potentially contributing to better health outcomes.

## Contributors

The study protocol was developed by JRG, JIT, AB, JM, TP, PP, and BH. Study implementation was managed by BK, MS, PN, VK, CK, MRR, MM, and BH. Study data was managed by BK, and MS while analysis was conducted by BK and CK. The initial draft of this manuscript was prepared by BK and reviewed by JIT, CPC, MRR and PP. Subsequent drafts were reviewed by JRG, BH, IB and PP. All authors proofread and approved the final version of the manuscript.

## Patient and public involvement

A cohort of influential traditional leaders in Eastern Province and three Chadiza-based traditional leaders were engaged through introductory meetings that covered the study objectives and core processes. At these meetings, each stage of the protocol was summarily discussed and subjected to participant input to ensure alignment with community needs and pave the way for later community engagement. During the process, minor refinements to the original protocol were necessary. Following community leader engagement, school system managers from across Chadiza, who normally convey community news, were engaged to galvanize family discourse around study processes. Community health workers (CHWs) were then familiarized with the study objectives and design, as well as their role, which they shared with their catchment communities. This sharing of information by CHWs with the families they served generated community buy-in, enabling the seamless implementation of weekly household visits by the same CHWs. Localized sharing of study processes formed the basis for strong community ownership, as community members not only understood the phases of the study, from enumeration to dissemination, but were able to provide invaluable insights into how to continuously harness their participation.

## Ethics approval

Study approval was granted by the Zambian ERES-Converge Research Ethics Committee (Ref no: 2020-Jan-003), the National Health Research Authority of Zambia, WCG IRB (IRB Protocol #20210568), and the Centers for Disease Control and Prevention IRB (7278).

## Trial registration

NCT04839900

## DISCLAIMER

The author’s views expressed in this publication do not necessarily reflect the views of the United States Agency for International Development (USAID), the United States Centers for Disease Control and Prevention, the United States President’s Malaria Initiative (PMI), or the United States Government.

## Data Availability

The data that support the findings of this study are available upon request from the corresponding author. The data are not publicly available due to ethical restrictions, as they contain sensitive information or identifiers that could potentially lead to the identification of study participants.

## References

1. World Health Organization. World malaria report 2024. Geneva; 2024.

2. PATH MACEPA. ZambiaMalariaRapidReportingDashboards [Internet]. Tableau; 2024. Available from: https://tableau.path.org/#/site/MACEPA-Zambia/views/ZambiaMalariaRapidReportingDashboards/RiskOverview?:iid=1

3. Zambia Ministry of Health. Zambia National Malaria Indicator Survey 2021. Lusaka; 2021.

4. Johnson AD, Thomson DR, Atwood S, Alley I, Beckerman JL, Koné I, et al. Assessing Early Access to Care and Child Survival during a Health System Strengthening Intervention in Mali: A Repeated Cross Sectional Survey. Eisele T, editor. PLoS ONE. 2013;8:e81304.

5. Linn AM, Ndiaye Y, Hennessee I, Gaye S, Linn P, Nordstrom K, et al. Reduction in symptomatic malaria prevalence through proactive community treatment in rural Senegal. Trop Med Int Health. 2015;20:1438–46.

6. Gaye S, Kibler J, Ndiaye JL, Diouf MB, Linn A, Gueye AB, et al. Proactive community case management in Senegal 2014–2016: a case study in maximizing the impact of community case management of malaria. Malar J. 2020;19:166.

7. Ratovoson R, Garchitorena A, Kassie D, Ravelonarivo JA, Andrianaranjaka V, Razanatsiorimalala S, et al. Proactive community case management decreased malaria prevalence in rural Madagascar: results from a cluster randomized trial. BMC Med. 2022;20:322.

8. Johnson AD, Thiero O, Whidden C, Poudiougou B, Diakité D, Traoré F, et al. Proactive community case management and child survival in periurban Mali. BMJ Glob Health. 2018;3:e000634.

9. Echodu D, Wanzirah H, Hadley L, Elliott R, Colborn K, Eganyu T, et al. Comparative Effectiveness Trial of Two Community Case Management Techniques Following Withdrawal of Indoor Residual Spraying In Ne Uganda. 2019.

10. Rutagwera M-RI, Ferriss EL, Kabamba BM, Porter T, Kangale CC, Gallalee S, et al. Impact of proactive malaria community case management (proCCM) on parasite prevalence and incidence from 2021 to 2023: a randomised controlled trial in Chadiza District, Eastern Province, Zambia. BMJ Glob Health. 2025;10:e017697.

11. PATH. Pre-elimination Interim Assessment Report. Lusaka; 2022.

12. CommCare (Version 2.52) [Internet]. Dimagi; 2008. Available from: https://play.google.com/store/apps/details?id=org.commcare.dalvik&hl=en&gl=US

13. Bowen DJ, Kreuter M, Spring B, Cofta-Woerpel L, Linnan L, Weiner D, et al. How We Design Feasibility Studies. Am J Prev Med. 2009;36:452–7.

14. Chomczynski P. QDA MINER – The Mixed Method Solution for Qualitative Analysis by Provalis Research: Software Review. Qual Sociol Rev. 2008;4:126–9.

15. Box [Internet]. 2024. Available from: https://www.box.com

16. World Health Organisation. WHO guideline on health policy and system support to optimize community health worker programmes. Geneva: WHO;

17. Yeboah-Antwi K, Pilingana P, Macleod WB, Semrau K, Siazeele K, Kalesha P, et al. Community Case Management of Fever Due to Malaria and Pneumonia in Children Under Five in Zambia: A Cluster Randomized Controlled Trial. Whitty CJM, editor. PLoS Med. 2010;7:e1000340.

18. Whidden C, Kayentao K, Liu JX, Lee S, Keita Y, Diakité D, et al. Improving Community Health Worker performance by using a personalised feedback dashboard for supervision: a randomised controlled trial. J Glob Health. 2018;8:020418.

19. Abbas F, Monroe A, Kiware S, Khamis M, Serbantez N, Al- Mafazy A-W, et al. Stakeholder perspectives on a door-to-door intervention to increase community engagement for malaria elimination in Zanzibar. Malar J. 2023;22:51.

20. Angotti N, Bula A, Gaydosh L, Kimchi EZ, Thornton RL, Yeatman SE. Increasing the acceptability of HIV counseling and testing with three C’s: Convenience, confidentiality and credibility. Soc Sci Med. 2009;68:2263–70.

21. Ministry of Health National Malaria Elimination Centre. National Malaria Elimination Strategic Plan 2017-2021. Lusaka: Ministry of Health National Malaria Elimination Centre; 2017.

